# Equivalence of Freeze-dried and Liquid-frozen Formulations of MVA-BN as Smallpox and Mpox Vaccine

**DOI:** 10.1101/2024.03.21.24304540

**Authors:** Richard N Greenberg, Darja Schmidt, Daniela Reichhardt, Siegfried Roesch, Sanja Vidojkovic, Jane Maclennan, Liddy M Chen, Robert Gruenert, Christian Kreusel, Heinz Weidenthaler, Thomas PH Meyer, Paul Chaplin

## Abstract

Orthopox virus-induced diseases such as smallpox and mpox (also known as monkeypox previously) remain a serious public health concern. Modified Vaccinia Ankara Bavarian Nordic (MVA-BN) has been approved in its liquid-frozen (LF) formulation for prevention of smallpox and mpox in the US, Canada and EU. A freeze-dried (FD) formulation might confer additional benefits such as longer shelf life and less reliance on cold chain storage and transport, thus can better meet the potential challenge of large quantity vaccine deployment in emergency situations.

In a phase 2 clinical trial, 651 vaccinia-naïve participants were vaccinated with two doses of MVA-BN LF or FD, 4 weeks apart. The objectives were to compare MVA-BN FD with LF in terms of vaccine-induced immune responses, safety and reactogenicity.

Strong vaccinia-specific humoral and cellular immune responses were induced by both formulations, with peak humoral responses at Week 6 and peak cellular responses at Week 2. At Week 6, geometric means of total antibody titers were 1096 (95% CI 1013, 1186) from the FD group and 877 (95% CI 804, 956) from the LF group, achieving the primary endpoint of non-inferiority of MVA-BN FD compared to MVA-BN LF. At Week 2, geometric means of T cell spot forming units were 449 (95% CI 341, 590) from the FD group and 316 (95% CI 234, 427) from the LF group. Both formulations of MVA-BN were well tolerated, with similar unsolicited AEs and solicited systemic reactions in both groups but slightly higher local reactions in the FD group. No vaccine related serious adverse events (SAEs) or vaccine related AE of special interest were reported.

The FD formulation of MVA-BN was shown to be equivalent to the LF formulation in immunogenicity, and comparable safety findings were observed from both formulations.

Clinical Trial Registration: NCT01668537

**Highlights:** Equivalence of MVA-BN freeze-dried and liquid-frozen formulations in immunogenicity MVA-BN FD and MVA-BN LF are comparable in clinical safety and reactogenicity Peak T cell responses were observed 2 weeks after the first vaccination

## Introduction

The world was declared free of smallpox in May 1980 as a result of concerted global vaccination campaigns using vaccinia virus-based vaccines [1, 2]. Subsequently, vaccination programs stopped, resulting in most of the present population being unprotected against smallpox or other orthopox viruses as the vaccinia virus-based vaccines also provide protection against other related orthopox viruses such as mpox. While the re-occurrence of smallpox remains a risk [3, 4], major concerns have been shifted to infections caused by other orthopox viruses due to the recent global mpox outbreak.

What is striking is how the recent mpox outbreak is behaving differently from what was previously known of the disease [5]. Since the first cases of human mpox disease reported in the 1970s in Central Africa, cases had occurred mostly in the Democratic Republic of the Congo, the Republic of Congo, Nigeria, Liberia and Sierra Leone [6-9]. The first human mpox cases outside of Africa occurred 2003 when an outbreak was reported in the Midwest USA [10]. In late 2018 several mpox infections were reported in the UK, Israel and in Singapore in travelers from Nigeria, at least one leading to a secondary infection of a healthcare worker [11-14]. However, the 2022 outbreak affected many non-endemic countries and quickly became a Public Health Emergency of International Concern, as declared by the World Health Organization (WHO) in July, less than 3 months since cases were reported from countries where the disease is not endemic. The outbreak was different in many ways besides major mode of transmission being sexual or intimate contact, predominantly in men having sex with men. Many cases were found to have lesions at multiple stages of development which was previously considered “unusual” for mpox [5]. This leads to questions about how long mpox has been circulating in its present form and concerns about other potential emergent viruses that can be circulating, unrecognized before it attracts public awareness [15].

MVA-BN has been developed as a non-replicating smallpox vaccine in response to the safety concerns of traditional replicating smallpox vaccines. MVA-BN induced vaccinia-specific humoral and cellular immune responses, while at the same time establishing the vaccine’s favorable safety profile [16-26] compared to replicating smallpox vaccines [16, 21]. MVA-BN (liquid-frozen formulation) has been licensed as a smallpox and mpox vaccine in the EU (tradename IMVANEX^®^) and in the USA (tradename JYNNEOS^®^). In Canada, it is also licensed for immunization against related orthopox virus infections (tradename IMVAMUNE^®^). In response to the recent mpox outbreak, more than 1.8 million doses of MVA-BN have been administered. The effectiveness and safety of MVA-BN based on real-world evidence data have been summarized in several reports or studies [27-30].

Vaccines as liquid-frozen (LF) formulation must be kept frozen between production and point of use. Freeze-drying is a conservation process, that allows the product to be stored as a powder at higher temperatures and for longer times compared to the liquid or liquid frozen presentations. The process removes water by sublimation from sensitive, perishable materials by freezing the substance, then reducing the pressure and adding low heat to force the frozen water in the material to change directly from ice to vapor without melting. Therefore, the freeze-dried (FD) formulations are generally considered to be more stable with less strict temperature requirements [31, 32] and thus may provide a longer shelf life, as well as making them easier to transport and store. The developed MVA-BN FD formulation has a shelf life of 8 years at -20°C compared to the approved MVA-BN LF formulation that has a shelf life of 9 years at -80°C. This would enable a more rapid response to emergencies by having the ability to ship and distribute even to remote rural areas without the need for extreme cold freezers.

In this report we present data from a phase 2 clinical trial comparing the FD formulation of MVA-BN with LF MVA-BN for humoral and cellular immunogenicity and safety.

## Materials and Methods

### Trial Design and Participants

The clinical protocol was approved by institutional review boards and the trial was conducted in accordance with the Declaration of Helsinki and good clinical practice guidelines. The enrolled participants were informed of all trial aspects and signed an informed consent form. This randomized, double-blind, multicenter phase 2 clinical trial was conducted between 2013 and 2014 at 3 trial sites in the USA (clinicaltrials.gov registry number: NCT01668537). Healthy, male and female vaccinia-naïve participants (18 to 55 years of age) of any ethnic origin were eligible if they had a Body Mass Index (BMI) in the range ≥18.5 and <35 with no symptomatic cardiopulmonary or metabolic disease and no clinically significant laboratory values. Participants were randomly assigned (1:1) to receive 2 vaccinations (4 weeks apart) of either the LF or FD formulation of MVA-BN at a dose of 1 x 10^8^ Tissue Culture Infectious Dose 50% (TCID_50_). The primary objective of the trial was to assess non-inferiority of the humoral immune responses induced by the FD formulation of MVA-BN compared to MVA-BN LF by vaccinia-specific enzyme-linked immunosorbent assay (ELISA). The secondary objective was to assess and compare the MVA-BN FD formulation to the MVA-BN LF formulation in terms of safety and reactogenicity.

### Immunogenicity Assessments

Blood for immunogenicity testing was collected at baseline (week 0, before the first vaccination) and at weeks 2, 4, 6 and 8. After collection and processing, serum and peripheral blood mononuclear cell (PBMC) samples were cryopreserved until testing. Vaccinia-specific antibody responses were measured using a plaque reduction neutralization test (PRNT) and an ELISA on serum samples [17]. Vaccinia-specific T cell responses were measured using an interferon-γ (IFN-γ) enzyme-linked immunospot assay (ELISPOT) on PBMC samples. For individuals which were seronegative before vaccination, seroconversion was defined as the appearance of an antibody titer equal to or higher than the detection limit (2 for PRNT and 50 for ELISA). For individuals seropositive (detectable antibody titer) before vaccination, seroconversion was defined as an at least 2-fold increase in titer compared with the pre-vaccination titer. Analog definitions applied for the determination of T cell responses, with a detection limit of 50 for the ELISPOT. Testing was performed in accordance with GCLP guidelines.

### Safety Assessments

Participants were closely monitored on site for at least 30 minutes after each vaccination. Safety was monitored by performing physical examinations including vital signs, routine laboratory measurements (at screening and selected visits after first and second vaccination) and evaluating adverse events (AEs). Solicited and unsolicited symptoms were collected over 8- and 29-day periods post-vaccination, respectively, as well as any serious adverse events (SAEs) during the whole trial period. Solicited symptoms included injection site AEs (pain, erythema, swelling, induration, and pruritus) and systemic AEs (increased body temperature, headache, myalgia, chills, nausea, and fatigue) captured in a daily symptom diary and graded by severity. Additional AEs of special interest, defined as cardiac symptoms after vaccination, clinically significant electrocardiogram changes and elevated levels of troponin I, were monitored throughout the trial. Trial participants were monitored for safety until 6 months after the second vaccination.

### Statistical Methods

Humoral immune response and safety analyses were performed on the full analysis set, defined as participants who received at least 1 vaccination. Analyses of cellular immune responses were performed on the cellular set, a subset of participants in the full analysis set with PBMC samples collected. Immunogenicity endpoints were summarized by geometric mean titers (GMT; for PRNT and ELISA) or geometric mean spot forming units (GMSFU; for ELISPOT) and 95% confidence intervals (CI) that were calculated by taking the antilogarithm of the log_10_ titer or log_10_ SFU transformations of the means and the 95% confidence bounds. The log difference and its 95% CI were also converted to GMT ratios (LF/FD) by taking the antilogarithm. Non-inferiority would be declared if the upper bound of the 95% CI is ≤1.5. Antibody titers and SFU below the assay detection limit were given a value of “1” for calculation of GMT and GMSFU. Adverse events were evaluated per participant and by vaccination period.

### Vaccine and Vaccine Administration

For both formulations of MVA-BN (LF and FD), the bulk drug substance was produced according to cGMP standards at Bavarian Nordic (Kvistgård, Denmark), and filled, formulated and labeled at IDT Biologika GmbH (Dessau-Rosslau, Germany). The MVA-BN batches C00004 (MVA-BN LF) and C00007 (MVA-BN FD) were used.

MVA-BN LF was shipped and stored at -20°C ± 5°C and provided in liquid-frozen 0.5 mL aliquots with a nominal titer of 1 x 10^8^ TCID_50_ / dose. Besides live attenuated MVA-BN, each dose contained 0.61 mg Tris-hydroxymethyl-amino methane and 4.1 mg sodium chloride, with no preservatives or adjuvants. The FD formulation of MVA-BN was shipped and stored at 5°C ± 3°C and provided as lyophilized aliquots with a nominal titer of 1 x 10^8^ TCID_50_ / dose after reconstitution to 0.5 mL with water for injection. Besides live attenuated MVA-BN, the FD formulation contained the same excipients as the LF formulation plus DTSG stabilizer for freeze-drying (9.45 mg Dextran FP 40, 22.5 mg Sucrose and 0.054 mg L-glutamic acid monopotassium salt) without preservatives or adjuvants.

Participants received 2 subcutaneous vaccinations with 0.5 mL of MVA-BN LF or MVA-BN FD, 4 weeks apart, preferably in the non-dominant upper arm.

## Results

### Population

Of 1032 screened individuals, 380 were not randomized due to screen failures, unwillingness to participate or other reasons. 652 participants were allocated to the 2 trial groups, MVA-BN LF and MVA-BN FD. In the LF group 327 participants were vaccinated. In the FD group 1 participant, although randomized, declined participation, and 324 participants were vaccinated, therefore a total of 651 participants comprised the full analysis set (Figure 1). Most participants received both vaccinations (314 in the LF group and 315 in the FD group). For analysis of cellular immune response, PMBC samples were collected from 101 and 94 participants of the LF group and the FD group, respectively (cellular subset; total N=195). The 2 treatment groups were comparable for demographic and baseline characteristics. The mean age was 27.8 years in the LF and 27.6 years in the FD group. Both treatment groups had slightly more females (54.4% in the LF and 52.5% in the FD group) and participants were mostly White/Caucasian, followed by Black/African American (Table 1). The cellular subset had demographic data that were comparable between the groups, but some differences when compared to the full analysis set regarding race and ethnicity (Table 1).

**Figure 1.**
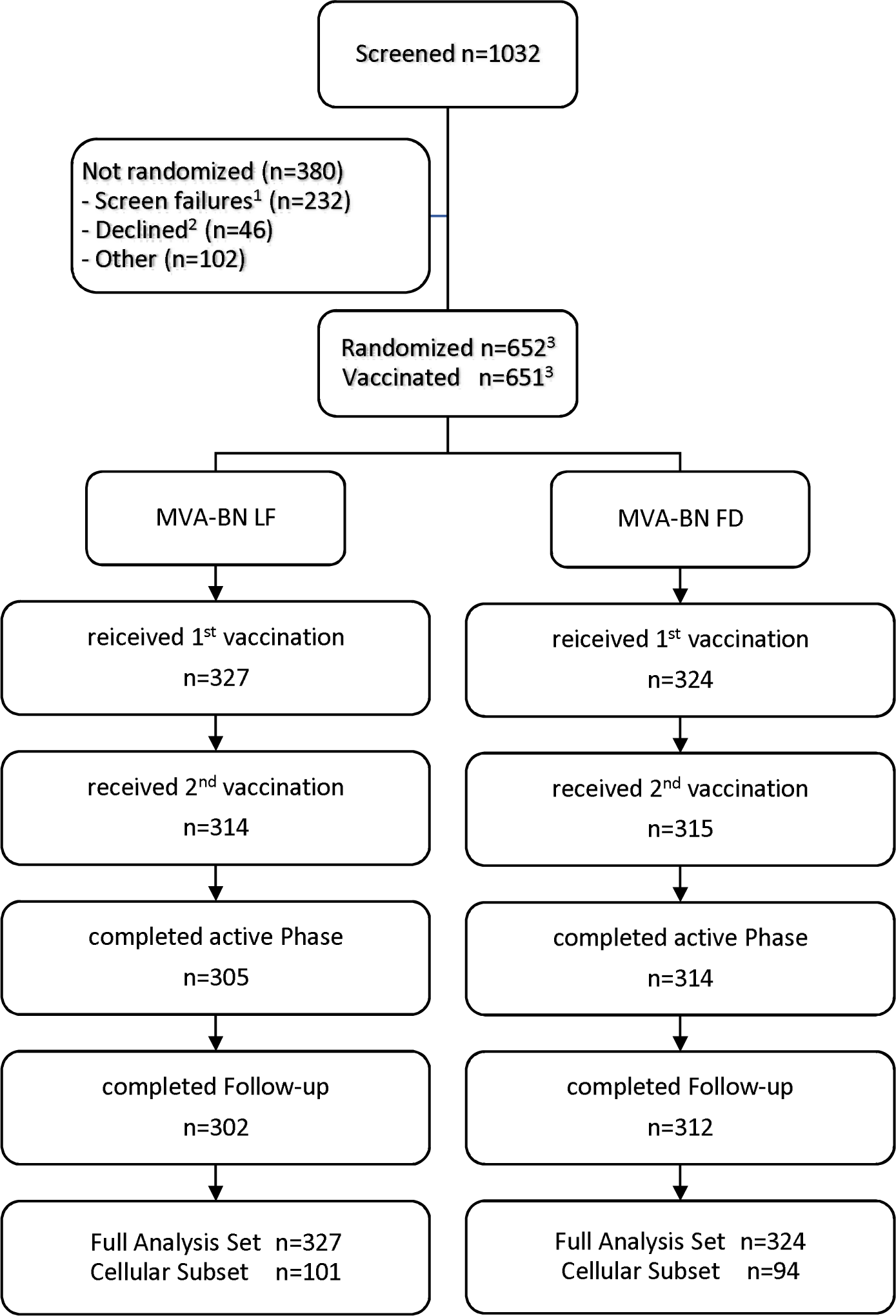
Disposition of Participants for Phase 2 Clinical Trial. Abbreviations: FD = freeze dried; LF = liquid frozen; n = number of participants per group. ^1^ not eligible ^2^ declined to participate ^3^ one individual declined to participate after randomization to group MVA-BN FD

**Table 1.** Demographic Data for Phase 2 Clinical Trial. Abbreviations: % = percentage based on N; BMI = body mass index; FD = freeze dried; LF = liquid frozen; N = number of participants; n = number of participants with data available; StD = standard deviation. ^1^ ‘Others’ includes American Indian/Alaskan Native, Asian, Native Hawaiian/Pacific Islanders, Other.

|  | **Full Analysis Set** |  | **Cellular Subset** |  |
| --- | --- | --- | --- | --- |
|  | **MVA-BN LF**<br>**(N=327)** | **MVA-BN FD**<br>**(N=324)** | **MVA-BN LF**<br>**(N=101)** | **MVA-BN FD**<br>**(N=94)** |
| **Age, years** |  |  |  |  |
| Mean (StD) | 27.8 (6.36) | 27.6 (6.21) | 30.4 (7.33) | 29.4 (6.72) |
| Median (range) | 27.0 (18, 55) | 27.0 (18, 47) | 29.0 (18, 55) | 29.0 (18, 47) |
| **Gender, n (%)** |  |  |  |  |
| Female | 178 (54.4) | 170 (52.5) | 57 (56.4) | 55 (58.5) |
| Male | 149 (45.6) | 154 (47.5) | 44 (43.6) | 39 (41.5) |
| **BMI [kg/m<sup>2</sup>]** |  |  |  |  |
| Mean (StD) | 25.64 (4.083) | 26.05 (4.190) | 26.58 (3.890) | 26.25 (4.230) |
| **Race, n (%)** |  |  |  |  |
| Black/African American | 78 (23.9) | 67 (20.7) | 37 (36.6) | 32 (34.0) |
| White/Caucasian | 239 (73.1) | 247 (76.2) | 62 (61.4) | 56 (59.6) |
| Others <sup>1</sup> | 10 (3.1) | 10 (3.1) | 2 (2.0) | 6 (6.4) |
| **Ethnicity, n (%)** |  |  |  |  |
| Hispanic or Latino | 95 (29.1) | 95 (29.3) | 3 (3.0) | 4 (4.3) |
| Non-Hisp. or Latino | 232 (70.9) | 229 (70.7) | 98 (97.0) | 90 (95.7) |

### Immunogenicity Results

As expected for a vaccinia-naïve population, most of the trial participants had no detectable vaccinia antibody titer prior to the first vaccination at week 0 (baseline), namely 98.5% and 97.8% were seronegative, as determined by PRNT and ELISA, respectively (data not shown). Vaccination with 2 doses of MVA-BN 4 weeks apart (complete vaccination schedule) resulted in rapid increases of PRNT and ELISA GMTs in both treatment groups with peak GMTs observed at week 6 (2 weeks after the second vaccination). PRNT and ELISA seroconversion rates were >85% after one vaccination (at week 4) and reached 100% in both groups at week 6 or 8. The PRNT GMTs at week 6 were higher in the FD group (GMT of 101.0; 95% CI: 91.0, 112.2) compared to the LF group (GMT of 83.3; 95% CI: 74.5, 93.1). A similar trend was observed for ELISA, with a GMT of 1095.9 (95% CI: 1012.6, 1186.0) in the FD group compared to 876.6 (95% CI: 803.7, 956.1) in the LF group (Figure 2, Table 2). The GMT ratio (LF/FD) of total antibodies measured by ELISA at week 6 was 0.800 (95% CI: 0.711, 0.899), demonstrating that the FD formulation induced antibody titers non-inferior and equivalent to those induced by the LF formulation, thereby meeting the trial’s primary endpoint. Similar to ELISA, the PRNT GMT ratios favored the FD formulation with a GMT ratio (LF/FD) of 0.824 (95% CI: 0.708, 0.960) at week 6 (Table 2).

**Figure 2.**
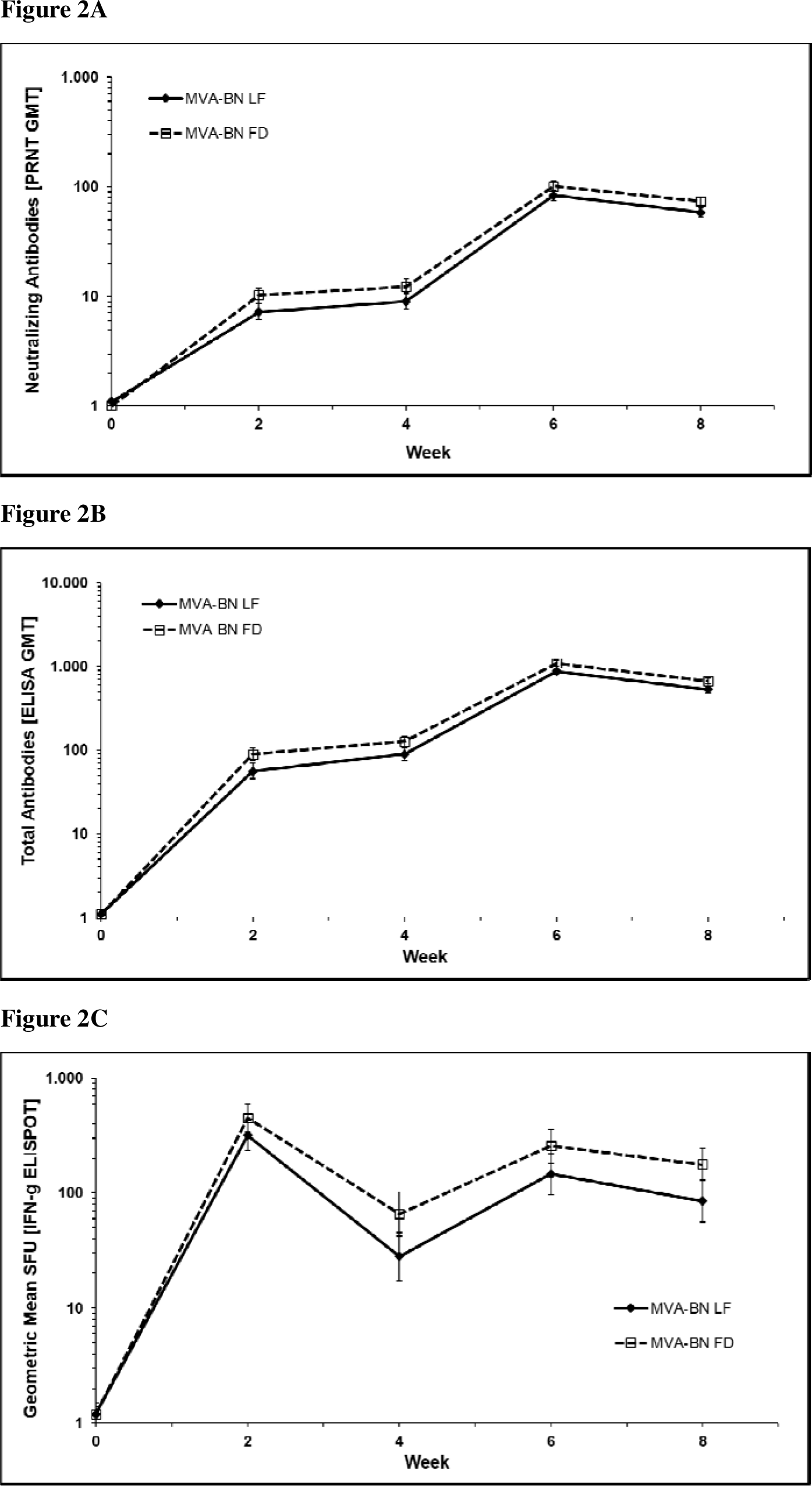
Humoral and Cellular Immune Responses for the Phase 2 Clinical Trial. Neutralizing antibody responses as determined by PRNT (**A**) and total antibody responses as determined by ELISA (**B**) in the full analysis set are shown as geometric mean titers (GMT) for MVA-BN LF and MVA-BN FD at all collection time points. (**C**) shows T cell responses as determined by IFN-γ ELISPOT in the cellular subset as geometric mean spot forming units (per 1 x 10^6^ PBMC) for MVA-BN LF and MVA-BN FD at all collection time points. Vaccinations were given at week 0 and week 4. Error bars are based on 95% confidence intervals. Abbreviations: ELISA = enzyme-linked immunosorbent assay; ELISPOT = enzyme-linked immuno spot technique; FD = freeze dried; GMT = geometric mean titer; IFN-γ = interferon gamma; LF = liquid frozen; PBMC = peripheral blood mononuclear cells; PRNT = plaque reduction neutralization test; SFU = spot forming units.

**Table 2.**
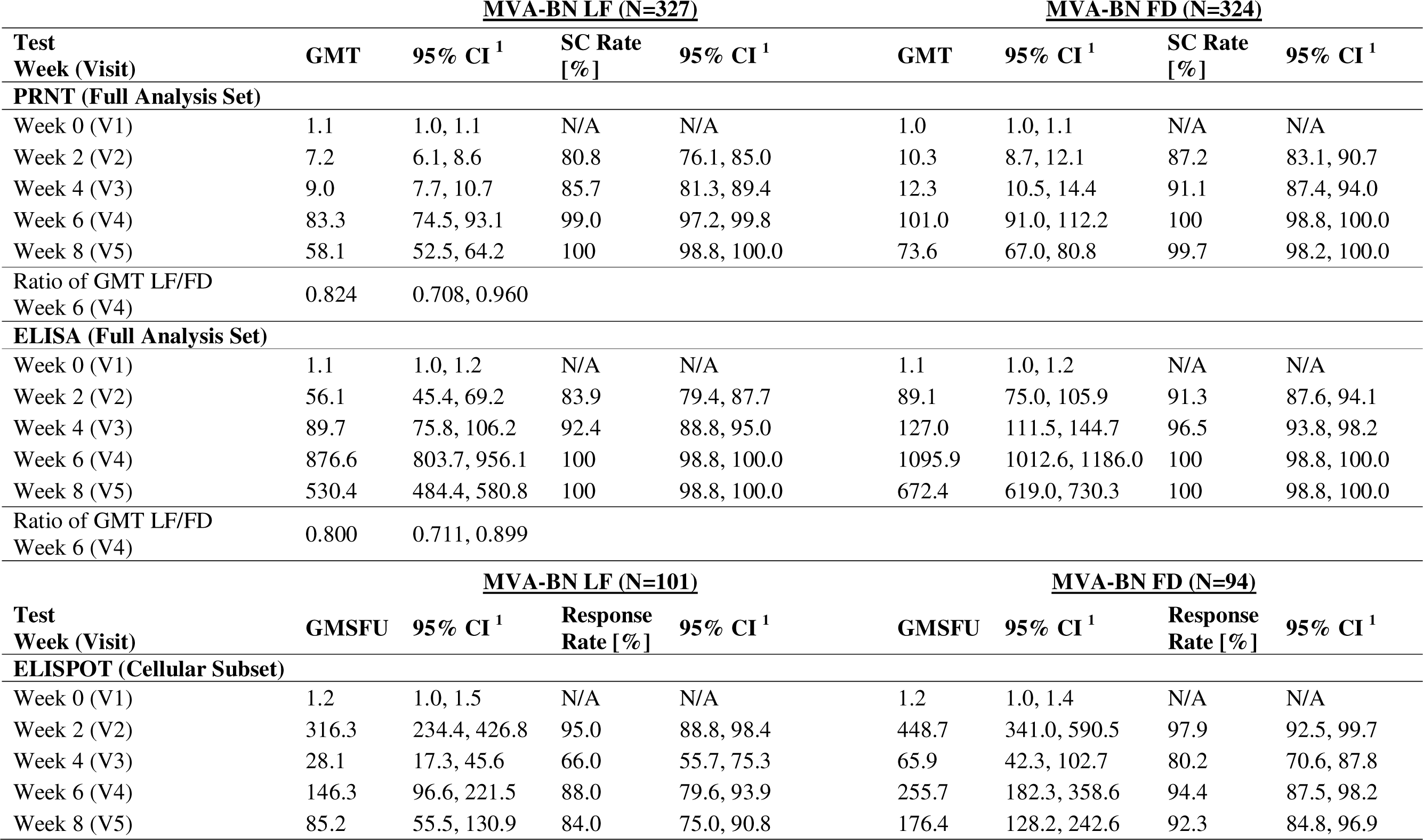
Humoral and Cellular Immune Responses in Participants of the Phase 2 Clinical Trial. Vaccinations were given at week 0 and at week 4. Abbreviations: CI = confidence interval; ELISA = enzyme-linked immunosorbent assay; ELISPOT = enzyme-linked immuno spot technique; FD = Freeze Dried; GMSFU = geometric mean spot forming unit (per 1 x 10^6^ PBMC); GMT = geometric mean titer; LF = liquid frozen; N/A = not applicable; N = number of participants per group; PBMC = peripheral blood mononuclear cells; PRNT = plaque reduction neutralization test; SC = seroconversion; V = visit. ^1^normal approximation to the log10 transformations 95% confidence interval, lower limit and upper limit

Most of the trial participants had non-detectable vaccinia-specific T cell responses at baseline, namely 95.0% (LF group) and 96.8% (FD group) as determined by IFN-γ ELISPOT (data not shown). Two weeks after the first vaccination, GMSFU had increased to 316.3 (95% CI: 234.4, 426.8) in the LF group and 448.7 (95% CI: 341.0, 590.5) in the FD group. GMSFU declined until week 4 and, following the second vaccination, increased again until week 6, although not as high as at week 2 (Figure 2, Table 2). Peak IFN-γ ELISPOT response rates were observed 2 weeks after the first vaccination (≥95%) in both groups and ranged from 66.0% to 94.4% from week 4 to week 8. Overall, there was no significant difference in T cell responses between the 2 vaccine formulations at any visit based on overlapping 95% CIs for GMSFU and response rates. All trial participants in both groups showed a T cell response to the vaccine at least at 1 post-baseline visit.

### Safety Results

The majority of participants in both groups experienced at least 1 adverse event (AE) during the clinical trial with 95.7% of the participants in both groups reporting at least 1 solicited or unsolicited AE (Table 3). The frequency of solicited local AEs was similar (91.7% of participants in the LF group and 93.2% in the FD group). Pain, erythema and swelling at the injection site were most frequently reported. For these local AEs participants from the FD group reported slightly more Grade 3 events compared to the LF group, namely: injection site pain (10.9% vs 6.8%), injection site erythema (14.6% vs 5.0%) and injection site swelling (7.2% vs 3.1%) (Table 4). The overall occurrence of solicited systemic AEs was comparable in both groups (55.7% in the LF group and 57.1% in the FD group) (Table 3); fatigue and headache were the most frequent solicited systemic AEs (Table 4). Grade ≥3 unsolicited AEs occurred at similar, low frequency in both groups: in 6 participants (1.8%) in the LF group and 5 participants (1.5%) in the FD group (Table 3); vomiting in 1 participant (FD group) was the only Grade 3 unsolicited AE reported by the investigator as causally related to the trial vaccine.

**Table 3.** Summary of Adverse Events for the Phase 2 Clinical Trial (Full Analysis Set) Abbreviations: % = percentage based on N; FD = freeze dried; LF = liquid frozen; N = number of participants with at least one documented vaccination period; n = number of participants with at least one event in any vaccination period. ^1^ Related: unsolicited or systemic solicited AE the investigator considered to have possible, probable, definite or missing relationship to trial vaccine; although solicited local events are by definition always considered related, they are not included here. ^2^ Serious adverse events are included even if they exceeded the 28-day follow-up period after each vaccination.

| **Safety Endpoint**<br>**Subject based** | **MVA-BN LF**<br>**(N=327)**<br>**n (%)** | **MVA-BN FD**<br>**(N=324)**<br>**n (%)** | **All Participants**<br>**(N=651)**<br>**n (%)** |
| --- | --- | --- | --- |
| Adverse Event | 313 (95.7) | 310 (95.7) | 623 (95.7) |
| Related <sup>1</sup> Adverse Event | 207 (63.3) | 207 (63.9) | 414 (63.6) |
| Unsolicited Adverse Event | 153 (46.8) | 157 (48.5) | 310 (47.6) |
| Unsolicited Adverse Event Graded<br>≥ 3 | 6 (1.8) | 5 (1.5) | 11 (1.7) |
| Unsolicited Related <sup>1</sup> Adverse Event<br>Graded ≥ 3 | 0 | 1 (0.3) | 1 (0.2) |
| Solicited Local Adverse Event | 300 (91.7) | 302 (93.2) | 602 (92.5) |
| Solicited Systemic Adverse Event | 182 (55.7) | 185 (57.1) | 367 (56.4) |
| Adverse Event of Special Interest | 1 (0.3) | 4 (1.2) | 5 (0.8) |
| Related <sup>1</sup> Adverse Event of Special<br>Interest | 0 | 0 | 0 |
| Serious Adverse Event <sup>2</sup> | 5 (1.5) | 2 (0.6) | 7 (1.1) |
| Related <sup>1</sup> Serious Adverse Event <sup>2</sup> | 0 | 0 | 0 |
| Deaths | 0 | 0 | 0 |
| Adverse Event leading to withdrawal<br>from vaccination | 0 | 1 (0.3) | 1 (0.2) |
| Adverse Event leading to withdrawal<br>from trial | 0 | 0 | 0 |

**Table 4.** Solicited Local and Systemic Adverse Events for the Phase 2 Clinical Trial **(Full Analysis Set)** Abbreviations: % = percentage based on N; FD = Freeze dried; LF = Liquid frozen; N = Number of participants with at least one memory aid completed; n = Number of participants with at least one event in any vaccination period. Related: adverse events the investigator considered to have possible, probable or definite relationship to the trial vaccine; or relationship was missing. All solicited local AEs were considered related to the trial vaccine.

|  | **MVA-BN LF**<br>**(N=322)**<br>**n (%)** | **MVA-BN FD**<br>**(N=321)**<br>**n (%)** | **All Participants**<br>**(N=643)**<br>**n (%)** |
| --- | --- | --- | --- |
| **Solicited Local Adverse Events** |  |  |  |
| Injection site pain |  |  |  |
| All Grades | 274 (85.1) | 290 (90.3) | 564 (87.7) |
| Grade 3 | 22 (6.8) | 35 (10.9) | 57 (8.9) |
| Injection site erythema |  |  |  |
| All Grades | 247 (76.7) | 255 (79.4) | 502 (78.1) |
| Grade 3 | 16 (5.0) | 47 (14.6) | 63 (9.8) |
| Injection site swelling |  |  |  |
| All Grades | 210 (65.2) | 223 (69.5) | 433 (67.3) |
| Grade 3 | 10 (3.1) | 23 (7.2) | 33 (5.1) |
| Injection site induration |  |  |  |
| All Grades | 211 (65.5) | 213 (66.4) | 424 (65.9) |
| Grade 3 | 5 (1.6) | 6 (1.9) | 11 (1.7) |
| Injection site pruritus |  |  |  |
| All Grades | 177 (55.0) | 189 (58.9) | 366 (56.9) |
| Grade 3 | 7 (2.2) | 11 (3.4) | 18 (2.8) |
| **Solicited Systemic Adverse Events** |  |  |  |
| Fatigue |  |  |  |
| All Grades | 113 (35.1) | 102 (31.8) | 215 (33.4) |
| Grade 3 | 15 (4.7) | 9 (2.8) | 24 (3.7) |
| Related Grade 3 | 14 (4.3) | 9 (2.8) | 23 (3.6) |
| Headache |  |  |  |
| All Grades | 125 (38.8) | 112 (34.9) | 237 (36.9) |
| Grade 3 | 8 (2.5) | 7 (2.2) | 15 (2.3) |
| Related Grade 3 | 6 (1.9) | 6 (1.9) | 12 (1.9) |
| Myalgia |  |  |  |
| All Grades | 68 (21.1) | 59 (18.4) | 127 (19.8) |
| Grade 3 | 1 (0.3) | 5 (1.6) | 6 (0.9) |
| Related Grade 3 | 1 (0.3) | 3 (0.9) | 4 (0.6) |
| Nausea |  |  |  |
| All Grades | 57 (17.7) | 51 (15.9) | 108 (16.8) |
| Grade 3 | 4 (1.2) | 2 (0.6) | 6 (0.9) |
| Related Grade 3 | 4 (1.2) | 2 (0.6) | 6 (0.9) |
| Chills |  |  |  |
| All Grades | 38 (11.8) | 35 (10.9) | 73 (11.4) |
| Grade 3 | 2 (0.6) | 5 (1.6) | 7 (1.1) |
| Related Grade 3 | 2 (0.6) | 4 (1.2) | 6 (0.9) |
| Body Temperature Increased |  |  |  |
| All Grades | 23 (7.1) | 33 (10.3) | 56 (8.7) |
| Grade $\geq 3$ | 3 (0.9) | 2 (0.6) | 5 (0.8) |
| Related Grade $\geq 3$ | 3 (0.9) | 2 (0.6) | 5 (0.8) |

Seven participants experienced a total of 7 serious AEs during the clinical trial: 5 in the LF group (spontaneous abortion, diabetic ketoacidosis, hemolytic anemia, limb injury, presyncope) and 2 in the FD group (anxiety, subcutaneous abscess). None of these events were considered related to the trial vaccine. No death occurred during the trial. There were no withdrawals from the trial due to AEs (Table 3). There were 6 adverse events of special interest reported in 5 participants (1 in the LF and 4 in the FD group), all events were of mild intensity and none of these events were considered related to the trial vaccination (Table 3). The most frequent adverse event of special interest was palpitations (1 event in 1 participant in the LF group and 3 events in 2 participants in the FD group). One participant reported tachycardia and another participant a second-degree atrioventricular block, both in the FD group after the first vaccination. No case of myo/pericarditis was reported.

## Discussion

Potential instability of a live viral vaccine can impose a considerable challenge to vaccine storage and distribution from manufacturing to the point of use [31-33]. Therefore, optimizing the product stability is an important goal for vaccine development. The SARS-CoV-2 pandemic has been a stress test for rapid deployment of vaccines that require extreme cold chain storage, especially in less developed countries. It is estimated that up to 80% of the cost of the vaccine is associated with the need for cold chain management [34, 35]. Drying of vaccine formulations provides one of the potential solutions to overcome these challenges [36, 37]. Bavarian Nordic developed a lyophilized formulation of MVA-BN with the aim to provide a smallpox/mpox vaccine that is suitable for long-term storage and can be easily deployed for immunization.

Immunogenicity and safety in this phase 2 clinical trial with 651 participants support the equivalence of both formulations. Both, total and neutralizing antibody titers, induced by MVA-BN FD were slightly higher than those from the LF group. Similar findings were also observed in a smaller NIH-sponsored phase 2 trial where an earlier lyophilized MVA-BN formulation was compared to MVA-BN LF [38]. However, these differences are not considered to be clinically meaningful, and the 2 formulations are deemed equivalent because the 95% confidence interval of the GMT ratio (LF/FD) falls between 0.67 and 1.5, the commonly acceptable equivalence limits for vaccines by regulatory agencies. It is possible that the thawing process and time between thawing and application can lead to minor degradation in the MVA-BN LF formulation. Although the roles of neutralizing antibodies have been well recognized, other types of immunity can also contribute to the control of viral infections and clearance. T cells have been shown to protect in the absence of neutralizing antibodies [39]. Antibodies against various conserved antigens of the viruses are generally non-neutralizing. However, their roles in exerting protective function through various immune mechanisms, such as signaling T cell responses, have been described [40-42]. In this study, besides demonstrating equivalence of T cell responses in a large number of participants, we observed that T cell responses reached peak signals at week 2 after a single vaccination while the increase after the 2^nd^ vaccination is smaller. This finding has significant implication because both the total antibody and the neutralizing antibody titers are still very low at week 2 and week 4 after a single vaccination. The critical role of T cell immunity in protection against orthopox viruses has been described in various publications [43-47] and may be responsible for long-lasting immunity against the viruses [44]. This finding leads to the belief that the vaccine may already offer protection against the viruses as early as 2 weeks after a single vaccination without the presence of antibodies. In a long-term follow-up and booster vaccination trial, it was found that even with nearly undetectable neutralizing antibody titers 2 years following initial MVA-BN vaccination, a MVA-BN booster induced robust anamnestic antibody responses, even stronger than peak responses following primary 1-dose or 2-dose vaccination [48]. These findings are consistent with data from observational studies that demonstrated 1-dose of MVA-BN showed 36% - 86% effectiveness [27, 28, 30, 49-51].

In addition to equivalent antibody immune responses, MVA-BN FD was comparable to MVA-BN LF in clinical safety. The incidences of Grade 3 injection site reactions were found to be slightly higher in the FD group compared to the LF group, possibly explained by higher total and neutralizing antibody titers after vaccination with MVA-BN FD. Such an association between vaccine adaptive immune responses and magnitudes of the inflammatory response has also been reported for other vaccines [52]. Unlike replicating smallpox vaccines which have contraindications for individuals with severe immunodeficiency [53], the non-replicating vaccine MVA-BN has a favorable safety profile without those contraindications [26, 54]. The favorable safety profile of MVA-BN is further supported by the findings from this trial. No vaccine related serious adverse event or cardiac inflammatory disorders were reported from the 651 participants.

The FD formulation used in this trial has been optimized with the same validated bulk drug substance process as for the LF formulation and was manufactured under the final industrial scale process. In a phase 3 clinical trial evaluating the lot-to-lot consistency of the FD formulation of MVA-BN, the 3 MVA-BN FD lots induced equivalent antibody responses with similar safety and reactogenicity reported from over 1000 participants [55].

## Conclusions

Equivalence of MVA-BN FD to MVA-BN LF was demonstrated in this phase 2 clinical trial. Both formulations of MVA-BN were well tolerated and further supported the favorable safety profile of MVA-BN. Findings from this trial, together with results from the phase 3 lot-to-lot consistency trial that demonstrated the consistency and reliability of the manufacturing process, support the application of MVA-BN FD as an alternative to MVA-BN LF.

## Data Availability

Anonymized data can be made available upon request. All requests must be submitted to Bavarian Nordic A/S for evaluation and approval.

## Acknowledgements

The study was funded by the Biomedical Advanced Research and Development Authority (BARDA) [grant number HHSO100201000011C]

